# Behavioural Activation for Social IsoLation (BASIL^+^) trial (Behavioural activation to mitigate depression and loneliness among older people with long-term conditions): Protocol for a fully-powered pragmatic randomised controlled trial

**DOI:** 10.1101/2022.01.29.22270065

**Authors:** BASIL+ trial collective, Lauren Burke, Elizabeth Littlewood, Samantha Gascoyne, Dean McMillan, Carolyn A. Chew-Graham, Della Bailey, Claire Sloan, Caroline Fairhurst, Kalpita Baird, Catherine Hewitt, Andrew Henry, Eloise Ryde, Leanne Shearsmith, Peter Coventry, Suzanne Crosland, Elizabeth Newbronner, Gemma Traviss-Turner, Rebecca Woodhouse, Andrew Clegg, Tom Gentry, Andrew Hill, Karina Lovell, Sarah Dexter Smith, Judith Webster, David Ekers, Simon Gilbody

## Abstract

**Introduction:** Depression is a leading mental health problem worldwide. People with long-term conditions are at increased risk of experiencing depression. The COVID-19 pandemic led to strict social restrictions being imposed across the UK population. Social isolation can have negative consequences on the physical and mental wellbeing of older adults. In the Behavioural Activation in Social IsoLation (BASIL^+^) trial we will test whether a brief psychological intervention (based on Behavioural Activation), delivered remotely, can mitigate depression and loneliness in older adults with long-term conditions during isolation.

**Methods:** We will conduct a two-arm, parallel-group, randomised controlled trial across several research sites, to evaluate the clinical and cost-effectiveness of the BASIL^+^ intervention. Participants will be recruited via participating general practices across England and Wales. Participants must be aged ≥65 with two or more long-term conditions, or a condition that may indicate they are within a ‘clinically extremely vulnerable’ group in relation to COVID-19, and have scored ≥5 on the Patient Health Questionnaire (PHQ9), to be eligible for inclusion. Randomisation will be 1:1, stratified by research site. Intervention participants will receive up to eight intervention sessions delivered remotely by trained BASIL^+^ Support Workers and supported by a self-help booklet. Control participants will receive usual care, with additional signposting to reputable sources of self-help and information, including advice on keeping mentally and physically well.

A qualitative process evaluation will also be undertaken to explore the acceptability of the BASIL^+^ intervention, as well as barriers and enablers to integrating the intervention into participants’ existing health and care support, and the impact of the intervention on participants’ mood and general wellbeing in the context of the COVID-19 restrictions. Semi-structured interviews will be conducted with intervention participants, participant’s caregivers/supportive others and BASIL^+^ Support Workers.

Outcome data will be collected at one, three, and 12 months post-randomisation. Clinical and cost-effectiveness will be evaluated. The primary outcome is depressive symptoms at the three-month follow up, measured by the PHQ9. Secondary outcomes include loneliness, social isolation, anxiety, quality of life, and a bespoke health services use questionnaire.

**Discussion:** This study is the first large-scale trial evaluating a brief Behavioural Activation intervention in this population, and builds upon the results of a successful external pilot trial.

**Trial Registration:** ClinicalTrials.Gov identifier ISRCTN63034289, registered on 5th February 2021.

**Funding:** This study was funded by National Institute for Health Research (NIHR) Programme Grants for Applied Research (PGfAR) RP-PG-0217-20006

## Introduction

Long-term health conditions (LTCs) are reported to be more prevalent in older people, and numbers continue to increase. This includes the number of older people with multiple LTCs [1]. The co-existence of two or more chronic conditions is termed ‘multiple long-term conditions’ (MLTCs) by the National Institute for Health Research (NIHR) [2]. NICE guidance highlights the importance of individualised treatment plans that address both physical and psychological conditions [3].

Depression is a major contributor to the overall global burden of disease according to the World Health Organisation [4] and is set to become the highest among all health problems by 2030 [5]. A two-to-three-fold increase in the prevalence of depression is found across the range of LTCs, resulting in poorer outcomes, lower quality of life and increased mortality [6]. Additionally, people with depression are more likely to have two or more LTCs and experience the greatest reduction in quality of life [7]. This results in substantially increased treatment costs and a significant contribution to health inequalities, particularly among older adults [8].

The Coronavirus 2 (SARS-CoV-2) virus (and the COVID-19 disease it causes) reached pandemic status in March 2020 triggering a national Stay At Home order (“lockdown”), where the UK government and devolved nations instructed individuals to follow compulsory social distancing and self-isolation rules. In addition, more than 2.2 million people were identified by the NHS as ‘clinically extremely vulnerable’ (including those with specific health conditions and older people) and were advised to follow stricter forms of isolation (“shielding”) [9]. Restriction levels have varied throughout the pandemic, with many people choosing to continue shielding, or maintain previous social distancing guidance, regardless of restrictions being eased [10].

The population’s mental health has worsened as a consequence of the pandemic [11], with reports of an increase in feelings of depression, anxiety and loneliness in older adults [12]. The social restrictions imposed to attempt to contain the virus led to widespread social and economic disruption. This rapid change to social norms increased isolation leading to loneliness and depression, which are known factors in the development of poorer mental health outcomes [12]. Existing research supports these reports, highlighting how social isolation has a disproportionate effect on the physical and mental wellbeing of older adults [13, 14]. Loneliness is also considered a significant public health concern in those aged 60 years and over, with evidence showing a link between persistent loneliness and increased healthcare utilisation, regardless of health status [13]. Furthermore, a longitudinal study of ageing indicated loneliness is linked to future depression, as depressive symptoms increased over time among people with higher loneliness scores [15]. Loneliness is not an inevitable consequence of social isolation, and approaches to mitigate loneliness were highlighted as population priority even before the COVID-19 pandemic [16, 17]. Such approaches include focusing on social networks [18] or adapting strategies derived from cognitive behavioural therapy in order to maintain social connectedness [19]. This emphasises the need for an intervention incorporating both depression and loneliness, which could potentially mitigate the immediate and longer lasting psychological impacts of COVID-19 on vulnerable populations, including older people and those with LTCs [20].

Behavioural Activation (BA) is a clinically effective structured psychological treatment for depression, encouraging individuals to reconnect with valued activities and positive reinforcement in their environment [21]. The therapist (or ‘case manager’) and patient work together to reinstate (or replace, if former activities are no longer possible) behaviours that improve mood and encourage the patient to remain active. It has the potential to address the disrupted routines and reduced opportunities for physical activity and social contact that have resulted from the increased social isolation older adults have faced over the course of the pandemic, whilst also addressing the social isolation that some older adults may have faced irrespective of the pandemic. BA is delivered within a Collaborative Care framework, which is a way of organising care so that an intervention is delivered in the most effective, patient-centred way. A Cochrane review has emphasised the importance of Collaborative Care in relation to multiple long-term conditions [22]. Evidence supports Collaborative Care for people with depression particularly if it includes a structured psychological intervention [22]. Given the nature of the COVID-19 pandemic, it is crucial that such interventions are able to be delivered remotely. Evidence suggests it is feasible and acceptable to use remote interventions to mitigate the psychosocial consequences (including depression and loneliness) of COVID-19 [23].

Recent research has described the impacts of the pandemic on the population’s mental health [24, 25]. However, clinical trials to test interventions to prevent and alleviate the psychological impact of COVID-19 have been limited, with drug and vaccine development trials prioritised [26]. By late 2019, we had co-developed a brief and scalable psychological intervention (based on BA) as part of an existing programme of work (Multi-Morbidity in Older Adults with Depression Study, MODS (RP-PG-0217-20006)). The MODS intervention was designed to be delivered mostly by telephone (with a face to face contact for the first session) for older adults with MLTCs and depression. Plans to test the MODS intervention were halted in March 2020 due to the pandemic. In response to the pandemic and in an effort to mitigate the psychological impacts of social isolation on older adults with long-term health conditions, we drew on the accumulated knowledge from MODS to adapt the MODS BA intervention to facilitate remote delivery – and developed the BASIL (Behavioural Activation for Social IsoLation) programme of work.

An external pilot Randomised Controlled Trial (RCT) commenced in April 2020 (BASIL-C19) to test the feasibility and acceptability of the BASIL intervention and remote trial delivery. The pilot study indicated that BA was a plausible intervention to mitigate the psychological impacts of COVID-19 isolation for older adults and is acceptable and feasibly delivered remotely [23]. The clinical and cost-effectiveness of the BASIL intervention will now be tested in a fully-powered large-scale RCT (BASIL+).

## Materials and Methods

### Research Aims

The aim of the BASIL^+^ trial is to test the clinical and cost-effectiveness of a brief psychological intervention (BA) with embedded process and economic evaluations:

- Establish the clinical effectiveness of BA in preventing and mitigating depression and loneliness among older people with multimorbidity during isolation.
- Establish the cost effectiveness of BA in preventing and mitigating depression and loneliness among older people with multimorbidity during isolation.
- To explore barriers and facilitators to implementation, in healthcare services, of the BA intervention for older people with multiple LTCs during isolation.

### Design

The BASIL^+^ trial is a two arm, parallel-group, multicentre, randomised controlled trial with embedded qualitative process and economic evaluations. The trial also includes a methodological Study Within a Trial (SWAT) to evaluate the impact of a study infographic on participant recruitment and response rates. The schedule of enrolment, interventions and assessments is shown in Figure 1.

**Fig 1.**
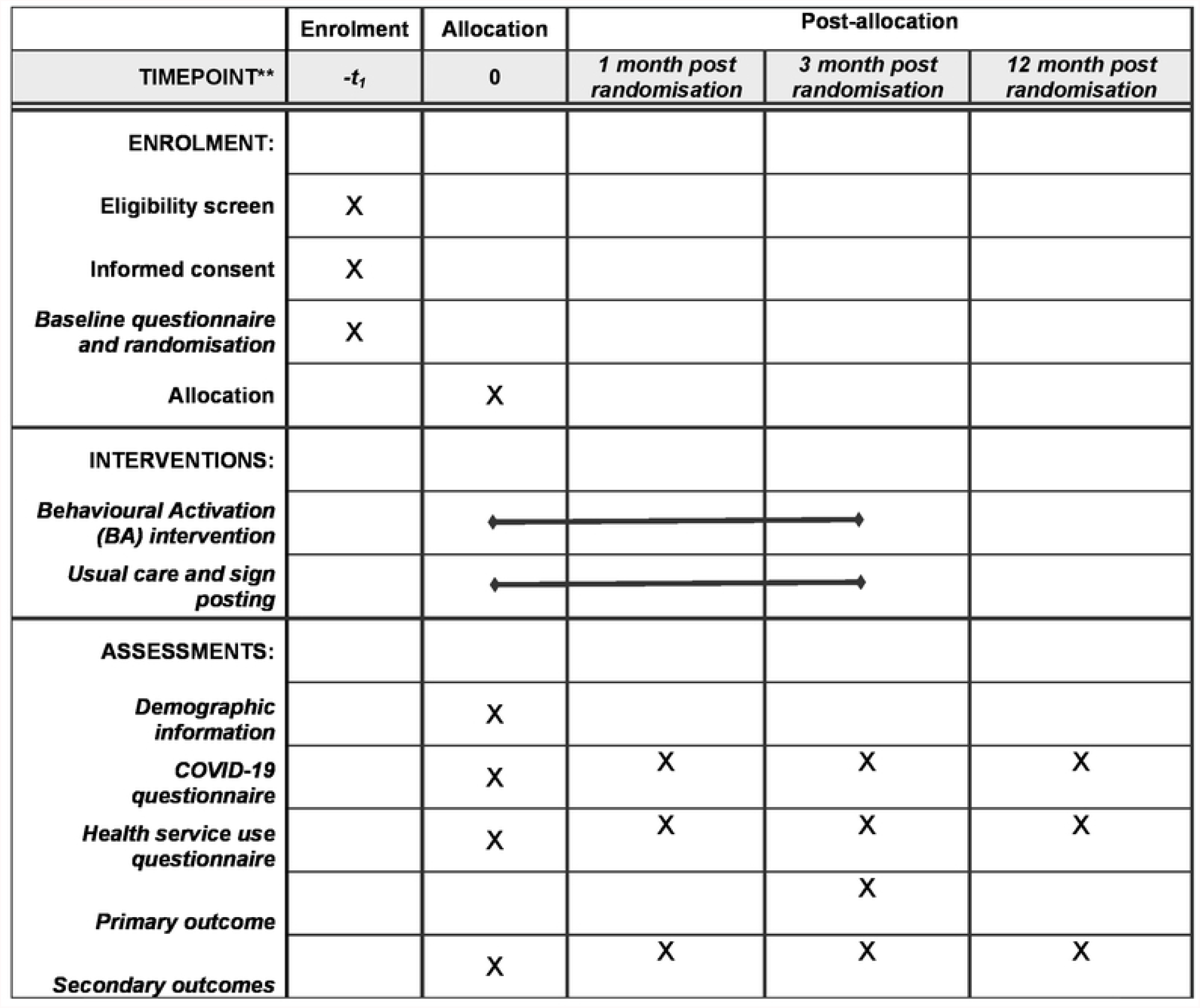
SPIRIT schedule of enrollment, interventions, and assessments.

### Setting

Participants will be identified via general practices across England and Wales. The intervention will be delivered across a range of health care settings (e.g. primary care, secondary care, voluntary/third sector).

### Identification

Potential participants will be identified via searches of patient registers at general practices. Lists of patients aged 65 and over with two or more long-term conditions (LTCs) or a condition that may indicate they are ‘clinically extremely vulnerable’ in relation to COVID-19 (following UK Health Security Agency (UKHSA), previously known as Public Health England guidance) will be generated. Patient lists will be screened by General practitioner (GP) practice teams to remove patients meeting exclusion criteria (detailed below).

LTCs will be based on the Department of Health (DoH) definition [27]. We will primarily focus on commonly reported LTCs for older people (such as asthma/ chronic obstructive pulmonary disease, diabetes, hypertension/coronary heart disease, and stroke) according to the primary care Quality and Outcomes Framework (QOF) [28] but will also include conditions such as musculoskeletal problems and chronic pain.

### Inclusion Criteria

- Older adults (65 years or over)
- Two or more long-term conditions (LTCs) or a condition that may suggest they are within a ‘clinically extremely vulnerable’ group in relation to COVID-19
- A score of ≥5 at screening on the Patient Health Questionnaire (PHQ9) (a score of 5-9 indicates possible sub-threshold depression, 10 and above indicates possible depression) [29]

### Exclusion Criteria

- Cognitive impairment
- Bipolar disorder/psychosis/psychotic symptoms
- Alcohol or drug dependence
- In the palliative phase of illness
- Have active suicidal ideation
- Currently receiving psychological therapy
- Unable to speak or understand English

Older adults will not be excluded on the basis of living in residential/care homes.

### Recruitment

Identified potential participants will receive a study information pack (containing a practice letter-headed invitation letter, Participant Information Sheet and an example consent form) and a text message about BASIL^+^ (where possible) via their GP practice. Patients will be given the option to complete an online consent form (via a link provided in the PIS, mirroring the example consent form patients receive), or to contact the study team directly. Extended GP practice teams will contact the remaining potential participants by telephone to determine interest in the study. Verbal permission to pass on their contact details (permission to contact (PTC)) to the local BASIL^+^ study team will be obtained from interested patients and recorded.

A BASIL^+^ study researcher will then make contact via telephone (or video call using an appropriate online platform) to discuss the study, answer any questions and determine eligibility. Where patients are eligible and wish to participate, informed consent will be obtained (where online consent has not already been provided). In response to the need to deliver the trial remotely, the informed consent process will involve taking consent verbally. Participants will be directed to the example consent form provided in the study information pack; the a researcher will read out each of the consent statements, and request that the participant respond to confirm whether they agree or disagree with the statement, and creating a written record of their responses. Once informed consent has been confirmed, the baseline questionnaire will then be completed over the telephone with a researcher (either immediately following confirmation of eligibility or at a later date, in line with participant preference, and ideally within one week).

### Randomisation and blinding

Following baseline questionnaire completion, eligible and consenting participants will be randomised 1:1 to either receive the intervention (BA) or usual care with signposting information. Randomisation will take place via a secure online randomisation service provided by the York Trials Unit (YTU). A YTU statistician not involved with participant recruitment will generate the allocation schedule, using block randomisation stratified by site with randomly varying block sizes. Participants will be informed of their group allocation by telephone (and confirmed by letter). The participant’s GP will be informed by letter of the participant’s group allocation; this will include information about the participant’s mood symptoms (with the participant’s consent).

Researchers completing follow-up questionnaires (outcome measures) with participants over the telephone will be blind to group allocation. Due to the nature of the intervention, it is not possible to keep participants or the BASIL^+^ Support Workers delivering the intervention blind to allocation.

### Intervention

#### BASIL^+^ intervention

The BASIL^+^ intervention uses BA set within a Collaborative Care framework. BA aims to help people stay connected with the world by helping them to maintain activities which they value. Where valued activities may no longer be possible, either in the short or long-term, BA prompts participants to think about alternative activities which fulfil a similar function and help them to remain active. Remaining active and staying connected with the world may benefit physical and emotional wellbeing, which could be particularly important where COVID-19 restrictions have disrupted usual routines and connections with the outside world. The BASIL Support Worker (BSW) and the participant will work together using a self-help workbook to develop an individualised treatment plan. The plan will take into account relevant COVID-19 guidance and aid in the establishment of new daily routines. Support will be provided to help participants use technology to maintain contact with those important to them or access services. The booklet also recognises that participants may be worried about the current situation due to COVID-19, either about the pandemic itself or recommencing activities they have not undertaken for some time. The BSW can work with the participant to identify simple strategies to manage worry and help them to recommence activities in a way which makes them feel most comfortable.

The Collaborative Care aspect of the intervention may involve the BSW supporting the participant to take a proactive role in managing their mood and long-term health conditions. The BSW will liaise with the participant’s GP or other professionals involved in the participant’s care, if appropriate. They may also provide information about relevant support services or organisations running activities relevant to the participant’s interests.

Participants will be offered up to eight intervention sessions, delivered by a trained BSW and supported by the self-help booklet. The self-help booklet has been developed over a number of similar studies; most recently during the BASIL-C19 pilot study [23], where it was adapted for use during the COVID-19 pandemic with help and feedback from older adults, BSWs and a range of stakeholders, including members of the existing MODS/BASIL Patient and Public Involvement (PPI) group. Further minor adaptations were made for BASIL^+^ following the BASIL-C19 pilot (including minor design and wording changes to the self-help booklet). All sessions will be delivered remotely, either over the telephone or by video call (where this is available and preferred) and according to participant preference. The first session will last approximately one hour with subsequent sessions lasting approximately 30 minutes.

The depression scale of the Depression Anxiety Stress Scale (DASS) [30] will be used as a within session depression symptom monitoring tool. The DASS is a widely used and simple to score monitoring tool with clear clinical cut off scores (non/mild/moderate/severe). DASS scores will be used to aid decision making by BSWs in conjunction with their BASIL^+^ clinical supervisor. Where risk or significant clinical deterioration is indicated the participant will be supported to access more formal healthcare interventions.

### BASIL^+^ Support Workers (BSWs)

BSWs will have a range of backgrounds (clinical and non-clinical). They will be required to complete bespoke intervention training (approximately 22 hours) facilitated by clinical members of the study team. Materials, including recorded presentations, role-play demonstration of sessions and a support worker treatment manual will be provided to the BSWs prior to three, two-hour online/video group training sessions. The training will cover the components of the BA intervention within the Collaborative Care framework, intervention delivery, and study procedures. BSWs will be required to pass a telephone-based bespoke competency assessment with a training facilitator before commencing delivery of the intervention. BSWs will receive regular supervision/support from a clinical member of the BASIL^+^ study team.

### Comparator

Participants allocated to the usual care with signposting information group will receive their usual care as provided by their current NHS and/or third sector providers. In addition, usual care (control) participants will be signposted to sources of self-help and information, including advice on how to keep mentally and physically well. Examples of such sources includes guidance from UK Health Security Agency (UKHSA), previously known as Public Health England [31], and Age UK [32].

### Outcome measures

Data will be obtained at baseline and one, three, and twelve months post-randomisation. Baseline data will be collected over the telephone with a researcher. Participants will be offered the option to complete follow-up questionnaires online, via a secure emailed link, or over the telephone with a researcher. A reminder system including emails and letters will be in place to use where appropriate.

The primary outcome will be self-reported depression severity (as measured by the PHQ9) [29] at three months post-randomisation.

Secondary outcomes will include depression (PHQ9) [28] at one and twelve months, and loneliness (De Jong Gierveld Scale – 11 items) [33]; social isolation (Lubben Social Network Scale - 6 items) [34]; anxiety (GAD7) [35]; and health related quality of life (SF-12v2 [36] and EQ-5D-3L [37]) at one, three and twelve months post-randomisation. A bespoke questionnaire will be used to collect health service use data at all time-points. Information regarding COVID-19 circumstances will be collected at all-time points.

Demographic information will be collected at baseline, to include age, LTC types/health condition(s), socio-economic status, ethnicity, education, cohabitation status, number of children.

### Data management plan

Data will be securely stored on University or NHS computers and in secure settings. For those research staff working remotely, these servers will be accessed through secure organisation VPNs. Where data will be stored at non-NHS sites, the process for secure data storage will be reviewed and approved by the trial Sponsor. All data will be stored securely in line with GCP and GDPR guidance. Access to participant data will be restricted according to researcher role. Participant confidentiality will be maintained at all times unless significant risk to self or others is identified.

### Study Within a Trial (SWAT)

Relying on a text-heavy PIS has become the norm for many trials. There is conflicting evidence that introducing infographics (visual representations of information) has the potential to improve the participant experience [38, 39, 40].

A methodological Study Within a Trial (SWAT) will be embedded within BASIL+ to evaluate the effectiveness of a single page infographic, to go alongside the PIS, on participant recruitment and response rates. The infographic displays a participant’s journey through the BASIL^+^ trial. The use of the infographic will be tested in a cluster RCT embedded within the BASIL^+^ trial. Study sites (n=12 approx.) will be randomised 1:1 using simple randomisation to either the SWAT intervention group (potential participants receive PIS as usual, and infographic) or the SWAT control group (potential participants receive PIS as usual, and no infographic). Generation of the SWAT allocation sequence will be undertaken independently by a statistician at YTU not involved with the BASIL^+^ recruitment process. Study sites will be blind to SWAT group allocation.

The primary outcome of the embedded SWAT will be the participant recruitment rate into the BASIL^+^ trial. Secondary outcomes will be i) participant response rate in terms of expressions of interest in BASIL+ (permission to contacts received, direct participant contacts, or participant completion of the online consent form – see recruitment section above), and ii) retention in BASIL+ as measured by response to the three month post-randomisation participant questionnaire.

The proportion of participants randomised into the BASIL^+^ trial will be compared between the SWAT groups using a logistic regression model, adjusting for trial site as a random effect. The secondary outcomes will be similarly analysed; the model for response rate at the three-month follow-up will also include a covariate for BASIL+ allocation (intervention/usual care/not randomised). A simple cost analysis will be conducted to estimate the cost per extra participant recruited associated with the infographic.

### Safety considerations

A risk management protocol will be followed during all participant contact. Risk assessment training and standard operating procedures will be provided to all researchers and BSWs. Clinical members of local study teams will support the assessment of risk and communication of risk information to a participant’s GP or emergency services, as deemed appropriate.

Serious adverse events and adverse events will be monitored for by all researchers and BSWs and reported in a timely manner.

### Sample size

For populations with case-level depression, a successful treatment outcome has been defined as 5 points on the PHQ9 [41]; however, this was calculated using distribution methods, which are not currently advocated [42], and does not incorporate our wider population of interest. In an older population with subthreshold depression a difference of 1.3 has been found to be clinically and cost-effective [43] with a standard deviation of 4, equating to a standardised effect size of 0.325. An effect size of 0.3 is assumed for this calculation, which is also in line with pooled standardised mean differences found in a meta-analysis of previous trials of collaborative care [44]. Assuming 90% power, 5% alpha, 0.3 effect size and 20% attrition (from mortality and loss to follow-up) the trial would need to randomise 590 participants. However, this calculation is very conservative. In the primary analysis, baseline PHQ9 score shall be included as a covariate, which will result in gains in power. Therefore, accounting for the correlation between baseline and outcome PHQ9 score can reduce the required sample size. In the external pilot trial, a correlation of 0.58 was observed between the PHQ9 score at baseline and three months amongst participants who scored ≥5 at baseline, and follow-up at three months was 90% [23]. Assuming a correlation of at least 0.5, 90% power, 5% alpha, 0.3 effect size and 10% attrition, 392 participants would need to be recruited and randomised into the trial. Calculations conducted using Stata v15.

### Statistical analyses

A detailed statistical analysis plan will be produced prior to data analysis and approved by the joint Programme Steering and Data Monitoring and Ethics Committee. Analysis will be conducted on an intention to treat basis, using two-sided statistical tests at the 5% significance level, using Stata v17 or later. The statistician will not be blinded to treatment allocation.

The flow of participants through the trial will be detailed in a CONSORT diagram [Figure 2]. The number of individuals screened, eligible and randomised will be presented, with reasons for non-participation provided where available. Intervention adherence will be recorded and reported, including the number of sessions completed. The number of participants withdrawing from the intervention and/or the trial and any reasons for withdrawal will be summarised by trial arm.

**Figure 2:**
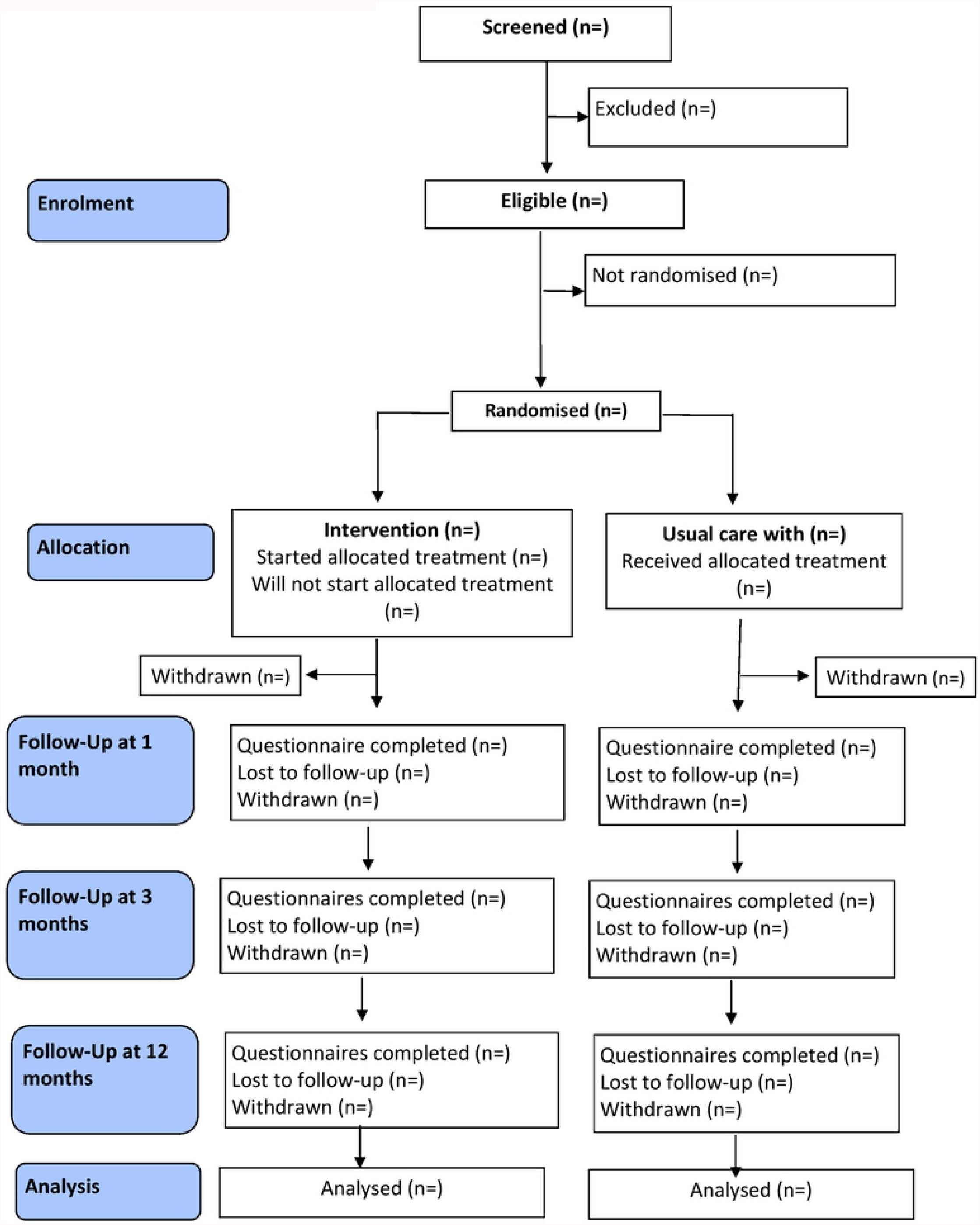
BASIL+ CONSORT flow diagram

Participant baseline data will be summarised descriptively by trial arm, for all randomised participants and for all who are included in the primary outcome analysis. No formal statistical comparisons will be undertaken on baseline data. Continuous measures will be reported as means and SD, while the categorical data will be reported as counts and percentages.

The primary outcome (PHQ9) will be analysed using a linear mixed model, incorporating data from all post-randomisation follow-up time points (one, three and 12 months). The model will include time, trial arm, an arm-by-time interaction, and baseline PHQ-9 score as fixed effects, with participant nested within site as random effects. The model will provide an overall treatment effect over time as well as estimates at individual time points, which will be reported as mean differences, 95% confidence intervals and p-values. The primary time-point of interest is pre-specified at three months.

The secondary outcomes of GAD7, De Jong Gierveld Scale, Lubben Social Network Scale, and SF-12v2 (mental and physical health component scores separately) will be similarly analysed.

A complier average causal effect (CACE) analysis will be conducted for the primary outcome to account for non-compliance with the intervention.

### Economic Evaluation

A trial-based economic evaluation from UK NHS and personal and social services (PSS) perspective will be conducted in order to evaluate the cost-effectiveness of the BA intervention compared to usual care for the target population. Health outcomes will be measured in terms of quality-adjusted life years (QALYs) using the EQ-5D-3L [37] questionnaire and calculated using standard area-under-the-curve method based on individual responses over the study period (45, 46). Costs of intervention and health service use will be calculated using a bottom-up costing approach. Resources used to deliver the intervention will be obtained from the study team, whereas health and social service resource use will be measured using a brief, tailored resource-use questionnaire, developed based on the questionnaire used in the BASIL C-19 pilot study [23]. Unit costs of health and social service use will be obtained from the UK national reference costs [47] and the Unit Costs of Health and Social Care report [48]. Cost of other professional support will be estimated from relevant salary scales and published reports/literature. Uncertainty around the incremental cost-effectiveness ratios (ICER) will be captured using non-parametric bootstrapping technique, and the results will be presented in the conventional form of a cost-effectiveness acceptability curve (CEAC). Sensitivity analyses will be conducted to assess the impact of using SF-12v2 to measure QALYs and the impact of including costs of informal care and other additional support.

### Qualitative process evaluation

The qualitative process evaluation will further explore the acceptability of the intervention, including important elements of the intervention and mode of delivery, barriers and enablers to integrating the intervention into participants’ existing health and care support, and the impact of the intervention on participants’ mood and general wellbeing in the context of the COVID-19 restrictions. Semi-structured interviews will be undertaken with samples of study participants, BSWs and caregivers/supportive others (where they have been involved in the intervention).

Interviews will be conducted with 15-20 participants who complete the intervention (“completers”), and up to 10 participants who were offered the intervention but who did not engage with or who ‘dropped out’ of the intervention. Up to 10 interviews will also be conducted with informal caregivers/supportive others. All participant and caregiver interviews will take place after the primary outcome has been completed.

Interviews will be conducted with up to 20 BSWs and will explore barriers and facilitators to delivery of the intervention and how the intervention might be integrated within health care settings. In order to assist with maximum variation in the experiences of the BSWs interviewed, factors such as intervention site location and demographic factors (such as job role, age and ethnicity) will be used to support selection for interview.

Interview topic guides will vary by participant group and will be modified iteratively as interviews and analyses progress. All interviews will last up to approximately 30-45 minutes and will be conducted over the telephone or a virtual platform.

Study participants provide consent to taking part in a qualitative interview as part of a set of optional consent statements (with form part of the trial consent form) upon entry to BASIL+. BSWs will obtain verbal permission from interested caregivers/supportive others for the BASIL^+^ study team to send them (post/email) a study information pack (containing an invitation letter, participant information sheet and example consent form) followed by telephone contact from the study team to discuss the study. For caregivers/supportive others who agree to take part in an interview, verbal consent will be taken (and audio recorded) prior to the interview.

BSWs will receive a study information pack (containing an invitation letter, information sheet and example consent form) following BSW training. They will be invited to complete an online consent form or to contact the qualitative study team directly to indicate their interest in taking part in an interview.

All interviews will be digitally-recorded (with participant consent), anonymised and transcribed, the transcripts forming the data for analysis. Verbal consent for interview participation will be taken (and recorded) before the interview commences. An initial thematic analysis will be conducted. Coding will be undertaken independently by members of the qualitative research team with meetings to ensure emerging codes remain grounded in the original data. A framework approach [49, 50] will be used to analyse the data, sensitised by the Normalisation Process Theory (NPT) [51] to explore barriers and facilitators to rapid implementation of the BASIL^+^ intervention into routine practice.

### Patient and Public Involvement

Our Patient and Public Involvement (PPI) Advisory Group (AG) was originally convened for the MODS programme of research and currently consists of a group (N=8) of individuals with a range of lived experience of physical and/or mental comorbidities and caregivers.

The PPI AG have extended their support to the BASIL research programme. Members of the research team have met virtually with the PPI AG on several occasions since the start of the COVID-19 pandemic. The PPI AG members provided feedback on the design and implementation of the BASIL+ trial, including the development of remote recruitment methods and materials (including the Participant Information Sheet) and plans for the remote delivery of the BASIL+ intervention and trial to include adherence to COVID-19 restrictions.

In addition to the work with the advisory group, the MODS/BASIL research programmes have PPI representation at Programme Management Group (PMG) meetings and on the independent Programme Steering and Data Monitoring and Ethics Committee (PSC).

### Status and timeline of the study

Recruitment of participants began in February 2021 and is expected to be completed by January 2022. Intervention delivery is expected to be completed by March/April 2022 depending on participant availability. The current protocol is version 4.0 dated 13/12/2021.

## Discussion

To the best of our knowledge, BASIL^+^ represents the first large-scale mental health randomised controlled trial conducted in response to the COVID-19 pandemic with the aim of mitigating the psychological impacts of the pandemic and associated social restrictions on potentially vulnerable older adults. The prevalence of depression and loneliness is expected to have increased in the general population due to COVID-19; this increase may have a disproportionate negative impact on older adults with existing long-term health conditions.

We adapted an evidence-based low intensity BA intervention, previously found to be effective at reducing depressive symptoms in older adults [52], for use during the pandemic. The brief nature of the intervention lends itself to remote delivery and could represent an efficient and effective way to support vulnerable populations by reducing depression and loneliness, particularly during periods of social isolation, such as the COVID-19 pandemic. The study has the potential to lead to significant benefits to the NHS, health and social care settings, and society.

BASIL^+^ will also add to existing trial based evidence in this area. There are few trials of cognitive or behavioural interventions [53], and we anticipate this will be the largest such trial. In time we anticipate that the results of BASIL^+^ will contribute to Cochrane reviews in this topic [54]. BASIL^+^ builds on the work conducted in the BASIL-C19 external pilot RCT [23] and will provide further lessons for the efficient delivery of mental health trials beyond the pandemic. The current work will be important in contributing to the emerging knowledge that trials can be conducted more efficiently and resourcefully when networks and research communities come together to work collaboratively with a united goal.

## Data Availability

No datasets were generated or analysed during the current study. All relevant data from this study will be made available upon study completion.

## Ethical considerations and declarations

The trial received ethical approval from Yorkshire and The Humber – Leeds West Research Ethics Committee on 11 December 2021 (REC Ref: 20/YH/0347). The sponsor for BASIL^+^ is Tees, Esk and Wear Valleys NHS Foundation Trust.

Although we are aware that the study population could be considered a vulnerable group, we do not anticipate any major ethical issues. The study will be conducted to protect the human rights and dignity of the participant as reflected in the 1996 version of the Helsinki Declaration. In order to protect the study participants, the following provisions will be made/upheld; the study has been designed to minimise pain, discomfort and fear and any foreseeable risk in relation to the experiences/treatments involved, the explicit wishes of the participant will be respected including the right to withdraw from the study at any time, the interest of the patient will prevail over those of science and society, provision will be made for indemnity by the investigator and sponsor. Patients will not be denied any form of care that is currently available in the NHS by participating in the study.

Protocol amendments will be managed via the Health Research Authority, Research Ethics Committee and sponsor approvals process during the study.

## Contributions of Authors

**Conceptualization:** Simon Gilbody, Elizabeth Littlewood, Dean McMillan, Carolyn A. Chew Graham, Della Bailey, Claire Sloan, Samantha Gascoyne, Peter Coventry, Catherine Hewitt, Gemma Traviss-Turner, Karina Lovell, Sarah Dexter Smith, Judith Webster, David Ekers, Andrew Clegg, Tom Gentry, Andrew J Hill, Karina Lovell

**Data curation:** Elizabeth Littlewood, Samantha Gascoyne, Lauren Burke, Catherine Hewitt, Rebecca Woodhouse, Elizabeth Newbronner, Caroline Fairhurst, Kalpita Baird, Leanne Shearsmith

**Formal analysis:** Caroline Fairhurst, Catherine Hewitt, Kalpita Baird, Elizabeth Newbronner, Carolyn A Chew-Graham, Peter Coventry, Leanne Shearsmith, Rebecca Woodhouse

**Funding acquisition:** Simon Gilbody, Elizabeth Littlewood, Carolyn A. Chew-Graham, Dean McMillan, Catherine Hewitt, David Ekers

**Investigation:** Andrew Henry, Eloise Ryde, Leanne Shearsmith, Dean McMillan, Elizabeth Newbronnner, Samantha Gascoyne, Lauren Burke, Rebecca Woodhouse, Della Bailey, Suzanne Crosland

**Methodology:** Elizabeth Littlewood, Carolyn A. Chew-Graham, Caroline Fairhurst, Catherine Hewitt, Gemma Traviss-Turner, Andrew Clegg, David Ekers, Andrew J Hill, Karina Lovell, Claire Sloan, Simon Gilbody, Dean McMillan, Peter Coventry

**Project administration:** Simon Gilbody, Dean McMillan, Lauren Burke, Elizabeth Littlewood, Samantha Gascoyne, Della Bailey, Suzanne Crosland, Carolyn A Chew-Graham, Peter Coventry, David Ekers

**Resources:** Dean McMillan, Della Bailey, Carolyn A Chew-Graham, David Ekers, Judith Webster, Claire Sloan

**Software:** Caroline Fairhurst, Catherine Hewitt, Kalpita Baird

**Supervision:** Simon Gilbody, Dean McMillan, Carolyn A. Chew-Graham, Elizabeth Littlewood, Della Bailey, Samantha Gascoyne, Suzanne Crosland, Rebecca Woodhouse, Lauren Burke

**Validation:** Caroline Fairhurst, Kalpita Baird, Catherine Hewitt, Simon Gilbody, David Ekers, Carolyn A Chew-Graham

**Visualization:** Lauren Burke, Samantha Gascoyne, Elizabeth Littlewood

**Writing – original draft:** Lauren Burke, Elizabeth Littlewood, Samantha Gascoyne, Rebecca Woodhouse, Andrew Henry, Eloise Ryde, Leanne Shearsmith, Kalpita Baird, Caroline Fairhurst, Simon Gilbody, Della Bailey

**Writing – review & editing:** Lauren Burke, Simon Gilbody, Dean McMillan, Carolyn A. Chew-Graham, Della Bailey, Samantha Gascoyne, Peter Coventry, Suzanne Crosland, Caroline Fairhurst, Andrew Henry, Catherine Hewitt, Kalpita Baird, Eloise Ryde, Leanne Shearsmith, Gemma Traviss-Turner, Rebecca Woodhouse, Andrew Clegg, Tom Gentry, Andrew J. Hill, Karina Lovell, Sarah Dexter Smith, Judith Webster, David Ekers, Elizabeth Newbronner, Claire Sloan

## Acknowledgements

We would like to thank: the participants for taking part in the trial, general practice and North East and North Cumbria Local Clinical Research Network staff for identifying and facilitating recruitment of participants, the independent Programme Steering Committee members for overseeing the study, and our PPI AG members for their insightful contributions and collaboration.

## Supporting Information

S1 File. SPIRIT checklist. (DOC)

S2 File. BASIL+ MT Trial Protocol v4.0 13.12.2021 (DOC)

